# Point of care lung ultrasound is useful when screening for CoVid-19 in Emergency Department patients

**DOI:** 10.1101/2020.06.09.20123836

**Authors:** Paul Walsh, Andrea Hankins, Heejung Bang

## Abstract

**Background:** CoVid-19 can be a life-threatening lung disease or a trivial upper respiratory infection depending on whether the alveoli are involved. Emergency department (ED) screening in symptomatic patients with normal vital signs is frequently limited to oro-nasopharyngeal swabs. We tested the null hypothesis that patients being screened for CoVid-19 in the ED with normal vital signs and without hypoxia would have a point-of-care lung ultrasound (LUS) consistent with CoVid-19 less than 2% of the time.

**Methods:** *Subjects:* Subjects were identified from ED ultrasound logs.

*Inclusion criteria:* Age 14 years or older with symptoms prompting ED screening for CoVid-19.

*Exclusion criteria:* Known congestive heart failure or other chronic lung condition likely to cause excessive B lines on LUS.

*Intervention:* Structured blinded ultrasound review and chart review

*Analysis:* We used a two-sided exact hypothesis test for binomial random variables. We also measured LUS diagnostic performance using computed tomography as the gold standard.

*Results:* We reviewed 77 charts; 49 met inclusion criteria. Vital signs were normal in 30/49 patients; 10 (33%) of these patients had LUS consistent with CoVid-19. We rejected the null hypothesis (p-value < 0.001). The treating physicians’ interpretation of their own point of care lung ultrasounds had a sensitivity of 100% (95% CI 75%, 100%) and specificity of 80% (95% CI 68%, 89%).

*Conclusion:* LUS has a meaningful detection rate for CoVid-19 in symptomatic ED patients with normal vital signs. We recommend at least LUS be used in addition to PCR testing when screening symptomatic ED patients for CoVid-19.

**Capsule:** *What is known:* Auscultation and chest x-ray are insufficient to screen for lung involvement when SARS-CoV-2 infection is suspected. Point of care lung ultrasound is widely available, safer, and less resource intensive than CT imaging.

*What we found:* In symptomatic patients presenting to the ED even those with normal vital signs had point of care lung ultrasound evidence of alveolar level involvement 33%of patients. Point of care lung ultrasound was 100% sensitive and 80% specific compared to CT (reference standard) when evaluating patients for Covid-19.

*What this adds:* Point of care lung ultrasound or similar imaging should performed when screening symptomatic patients in whom SARS-CoV-2 infection is suspected.

## Introduction

SARS-CoV-2 causes a variety of respiratory symptoms ranging from pharyngitis or rhinitis, through bronchitis, to multifocal peripheral pneumonitis extending to the alveoli. [1–3] Two clinically important characteristics of SARS-CoV-2 infection are that auscultatory findings may be subtle or normal even in the presence of advanced lower airway disease, and chest x-rays are inadequate for screening. [4] In common with other coronaviruses and influenza, SARS-CoV-2 is likely spread by both the droplet and airborne routes. [5–7] When aerosolized the resulting respirable particles less than 10μ in aerodynamic diameter contain viable virus and can reach adult alveoli directly. [8] Smaller aerosols (5μ) reach the alveoli without also being deposited in the bronchi.[8] This can lead to a clinical picture where a patient has serious lower respiratory tract infection with little or no concomitant upper respiratory tract infection. [6] Nasopharyngeal swabs, even if correctly collected, can therefore fail to detect SARS-CoV-2 and provide false reassurance despite ongoing alveolar destruction.

Screening for SARS-CoV-2 therefore frequently, but not always, includes both viral swabs from the oro-nasopharynx and imaging of the lower respiratory tract. This has included chest x-ray, computed tomography (CT) imaging, and sometimes point of care lung ultrasound. CT scan has been shown to be useful for screening and detecting patients with COVID-19 pneumonia, especially in the highly suspicious, a- or pre-symptomatic cases with negative nucleic acid testing. However, CT imaging is slow, exposes the patient to ionizing radiation, and exposes additional staff to SARS-CoV-2.[4, 9]

Point of care lung ultrasound can detect SARS-CoV-2 induced lung disease, is readily available in most emergency departments, (ED) does not expose the patient to ionizing radiation, and does not require the staff, expertise, and time necessary for traditional CT imaging.[10] Nonetheless, point of care lung ultrasound does add 5 to 10 minutes to the duration of patient evaluation, increases the treating physicians’ exposure to SARS-CoV-2, and decreases the number of patients seen hourly by that physician.

This raises the question as to whether lung imaging could be deferred if the patient being screened for SARS-CoV-2 has normal vital signs. Conversely, if the presence of normal vital signs does not preclude ultrasound evidence of lung disease then some current practices of swab only testing must be considered inadequate.

Our null hypothesis was that among symptomatic patients being screened for CoVid-19 in the emergency department that the point of care lung ultrasound would be consistent with CoVid-19 less than 2% of the time if vital signs were normal.

We also measured the diagnostic performance of point of care lung ultrasound compared with chest x-ray and CT chest. For comparative purposes we also measured the diagnostic performance of chest x-ray and crackles or rales on auscultation with CT chest.

## Methods

### Study Design

Cross sectional study with structured chart and ultrasound imaging review.

### Subjects

Patients aged 14 years old and older screened for CoVid-19 in an adult and pediatric emergency department were considered. Subjects were identified from the imaging archive of the ED ultrasound machine. Patients attended from 4-March-2020 to 19-May-2020. Patients had LUS performed if the treating physician was facile in point-of care LUS, believed that lung imaging should form part of CoVid-19 screening, and did not send the patient for immediate computed tomography of the chest. Although not randomly selected, the patients would not systematically differ from patients not imaged.

### Ultrasound imaging protocol

The physicians performing the point of care lung ultrasound typically imaged the posterior acoustic windows by running the US probe down the patient’s back midway between the scapula and vertebral column. Axillary and anterior windows were typically interrogated with single views of each. Physicians sometimes chose to not interrogate all possible windows if they had already reached their diagnosis on the windows already imaged. Images were captured with a Zonare Z One Ultra Ultrasound machine. The probes available for use were linear 10.5 MHz, linear 4.1 MHz, and curvilinear 9.3 MHz

### Inclusion Criteria

Images archived with adequate identifiers, age 14 years or older, and being evaluated for SARS-CoV-2 infection causing a CoVid-19 illness.

### Exclusion Criteria

Patients were excluded for a prior medical history of congestive heart failure or other chronic lung disease likely to affect point of care lung ultrasound interpretation (for example disease likely to cause B lines or pleural thickening) and if the point of care lung ultrasound was performed for a reason other than evaluating for CoVid-19. We did not exclude patients with a history of asthma or chronic obstructive pulmonary disease.

### Study definitions

We defined symptomatic as the documentation of any of the following in medical record: cough, subjective fever, fatigue, weakness, sore throat, or shortness of breath, nausea or vomiting, diarrhea, sore throat, fatigue, or headache.

Abnormal vital signs- Any pulse or respiratory rate at or above the 98th centile for age for children [11] For adults, tachycardia was defined as pulse at or above 100 beats per minute, tachypnea as respiratory rate above 22 breaths per minute, fever as temperature as at or above 38°Centigrade, and hypotension as systolic blood pressure at or below 80 mm Hg.[12] We did not have an upper limit for blood pressure. We included oxygen saturation measured by pulse oximetry as a vital sign and defined hypoxia as oxygen saturation of less than 92%.

We accepted the interpretation of the ultrasound by performing physician as consistent with CoVid-19 or viral pneumonitis for our primary analysis. If the performing physician did not document an interpretation we substituted the blinded reading.

### Data abstraction

One investigator (PW) performed a blinded reading of all LUS images using a structured template prior to performing chart review. Another, (AH) extracted data from EPIC/Clarity electronic health records using via SQL Server Management Studio.

Vital signs for each visit and polymerase chain reaction (PCR) results of swabs were extracted from their respective fields in an EPIC electronic health record. Only the first set of vital signs was retained. Vital signs and lab results were directly extracted from the electronic health record.

The full text of the ED visit was downloaded into a text file. EPIC electronic health record periodically automatically saves even incomplete notes as they are entered. The time of each, even incomplete note, is recorded. This allowed us to ensure the ultrasound note was entered before the CT scan resulted. However, the resulting downloaded files are duplicative and large (text fields alone average 1 megabyte per patient).

The ultrasound note was typically entered either in free-form or using personalized physician-created templates. These were in various locations in the chart. Some were typed into distinct stand-alone progress notes, others were included in the main chart, still others were included in progress notes that included other patient information. We used a simplified sentiment analysis (sentimentR) in R to locate the bedside ultrasound report in the chart.[13] This created an html page highlighting text that sentiment analysis considered to be an ultrasound report. In three cases a bedside ultrasound report could not be found using either this semi-automated technique or a manual chart review and the blinded interpretation was substituted.

CT scan and chest x-ray results have standardized headers and were located using regexm functions in Stata and then manually reviewed and data abstracted using a standardized template by an author (PW).

Because there was only one chart reviewer, inter rater reliability was not a concern. We did not attempt intra-rater reliability measurement of the chart abstraction process.

## Data and statistical analysis

We tested the null hypothesis using the bitest command in Stata. This performs exact hypothesis tests for binomial random variables. The null hypothesis was that the probability of a positive ultrasound was 2%. Our sample size calculations are shown in Appendix 1.

We compared inter rater reliability between the treating physician and reader relying on only the archived images using Gwet’s agreement coefficient (AC1). Gwet’s AC1’s validity does not depend upon the hypothesis of independence between raters and it does not result in unexpectedly low values (as seen in Cohen’s κ) when agreement is expected to be high.[14,15] We have previously shown how Cohen’s κ can be misleading in pediatric emergency medicine research and why alternatives such as Gwet’s AC1 should often be used instead.[16] We used kappaetc in Stata to calculate Gwet’ AC1. [17] We measured diagnostic performance of the point of care lung ultrasound using board certified radiologists’ interpretations of the CT chest as the gold standard using the diagt command in Stata. [18]

Data and statistical analysis was performed using Stata 16.1 (Statacorp LLP, College Station TX and R (R Foundation for Statistical Computing, Vienna, Austria).

## Ethical approval

The institutional review and privacy boards for Sutter Health approved this study and granted a waiver of informed consent (approval number 1597263).

## Results

We identified 77 point of care lung ultrasound scans with usable data of which 49 met our inclusion and exclusion criteria. All 77 scans were used to measure inter rater reliability and diagnostic performance characteristics. All the point of care lung ultrasounds were performed before the CT scans. Fig 1 shows patient flow through the study. The demographic characteristics of subjects are shown in Table 1.

**Table 1.** Clinical characteristics of study patients. IQR, interquartile range; LUS lung ultrasound.

|  |  | Total | LUS not suggestive of CoVid-19 | LUS suggestive of CoVid-19 |
| --- | --- | --- | --- | --- |
|  |  | N=49 | N=31 | N=18 |
| Gender | Male | 25(51%) | 13(42%) | 12 (67%) |
| Age (years) | Median (IQR) | 25 (15-46) | 22 (14-52) | 31 (16-46) |
| Duration (days) | Median (IQR) | 4 (2-7) | 3 (2-7) | 5(3-8) |
| Subjective fever at home | Present | 16 (33%) | 9 (29%) | 7 (39%) |
| Cough | Present | 26 (53%) | 15 (48%) | 11 (61%) |
| Dyspnea | Present | 29 (59%) | 18 (58%) | 11 (61%) |
| Sore throat | Present | 9 (18%) | 7 (23%) | 2 (11%) |
| Fatigue | Present | 9 (18%) | 6 (19%) | 3 (17%) |
| Headache | Present | 35 (71%) | 24 (77%) | 11 (61%) |
| Myalgias | Present | 5 (10%) | 4 (13%) | 1 (6%) |
| Diarrhea | Present | 6 (12%) | 2 (6%) | 4 (22%) |
| Nausea/vomiting | Present | 8 (16%) | 5 (16%) | 3 (17%) |
| Vital signs | Abnormal | 30 (61%) | 20 (65%) | 10 (56%) |
| Tachycardia | Tachycardia | 14 (29%) | 10 (32%) | 4 (22%) |
| Tachypneic | Tachypneic | 4 (8%) | 2 (6%) | 2 (11%) |
| Hypotension | Normotensive | 49 (100%) | 31 (100%) | 18 (100%) |
| Hypoxic | Hypoxia | 5 (10%) | 2 (6%) | 3 (17%) |
| Lungs clear on auscultation | Present | 35 (73%) | 23 (77%) | 12 (67%) |
| Crackles/Rales on auscultation | Present | 4 (8%) | 3 (10%) | 1 (6%) |
| Wheezing/ronchi on auscultation | Present | 6 (12%) | 3 (10%) | 3 (17%) |

**Figure 1.**
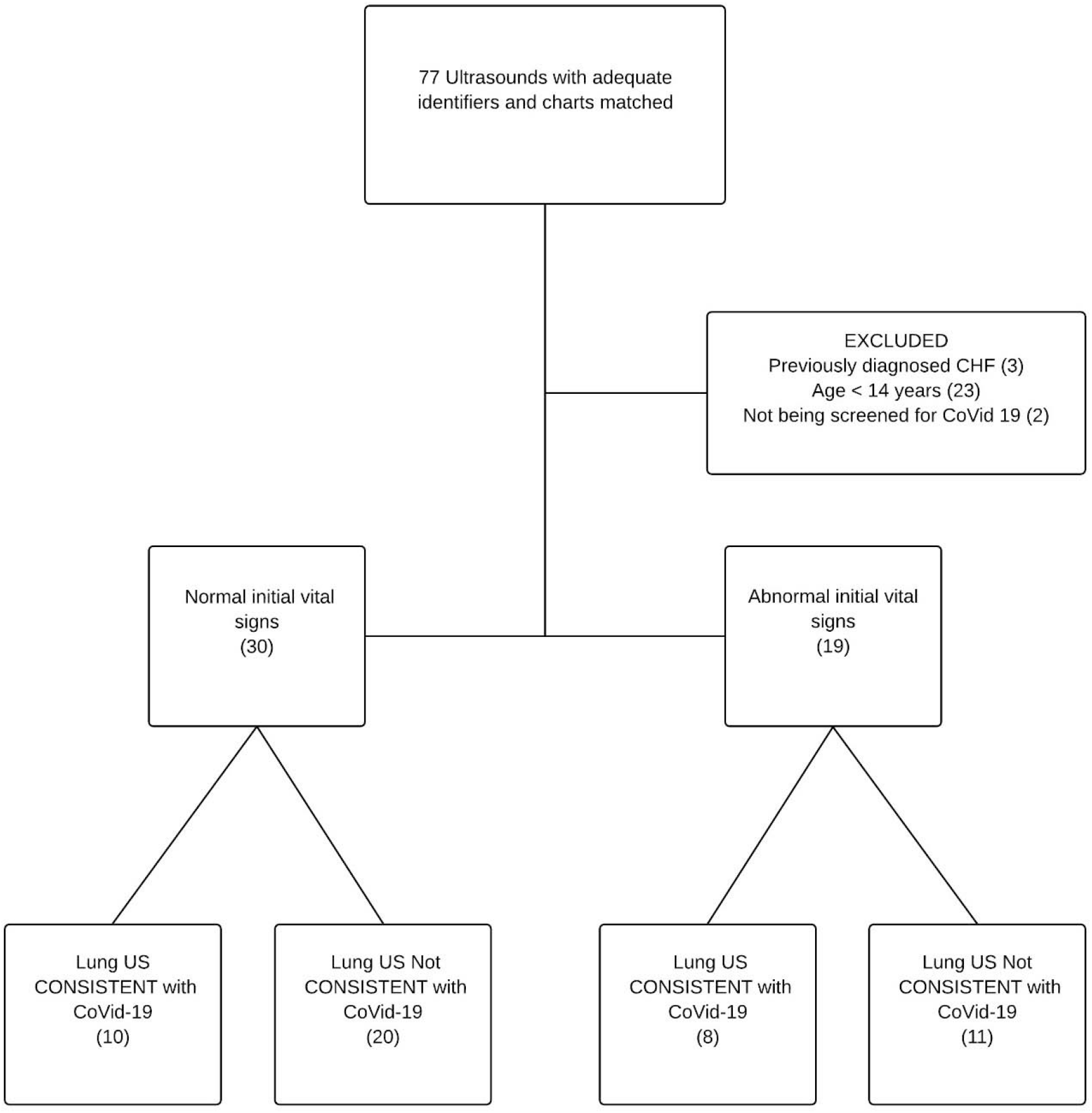
Patient flow through the study.

The treating physician interpreted 18/49 (37%) point of care lung ultrasounds as being consistent with Covid-19. Vital signs were normal in 30 patients and 10 (33%) of these patients had LUS consistent with CoVid-19.

We therefore reject the null hypothesis that among symptomatic patients being screened for CoVid-19 in the emergency department that the point of care lung ultrasound would be consistent with CoVid-19 less than 2% of the time if vital signs were normal (p-value < 0.001). We accept our alternative hypothesis that point of care lung ultrasound would be consistent with CoVid-19 more than 2% of the time even if the vital signs are normal.

When compared with the subsequent CT, the treating physicians’ interpretation of their own point of care lung ultrasounds had a sensitivity of 100% (95% CI 75%, 100%) and specificity of 80% (95% CI 68%, 89%). For the over reading physician relying only on archived images the sensitivity and specificity were 92% (95% CI 62%, 100%) and 37% (95% CI 25%, 50%) respectively. The performance characteristics of point of care lung ultrasound using CT chest as the gold standard are detailed in Table 2.

**Table 2.** Comparison of diagnostic performance of bedside point of care lung ultrasound, chest x ray, and crackles on auscultation for diagnosis of lung involvement of SARS-CoV-2 using CT chest as the gold standard. Predictive values were based on 16% prevalence. Sens, sensitivity; CI, confidence interval, Spec, specificity; PPV, positive predictive value; NPV, negative predictive value; LR +, likelihood ratio positive; LR –, likelihood ratio negative; AUC, area under the receiver operator curve.

| Modality | Sens | 95% CI | Spec | 95% CI | PPV | 95% CI | NPV | 95% CI | LR + | 95% CI | LR - | 95% CI | AUC | 95% CI |
| --- | --- | --- | --- | --- | --- | --- | --- | --- | --- | --- | --- | --- | --- | --- |
| Ultrasound | 100 | 75 100 | 80 | 68 89 | 48 | 28 69 | 100 | 93 100 | 4.7 | 2.9 7.7 | 0.1 | 0.0 0.7 | 0.90 | 0.85 0.95 |
| Chest x ray | 25 | 5 57 | 91 | 81 97 | 33 | 7 70 | 87 | 76 94 | 2.7 | 0.8 9.4 | 0.8 | 0.6 1.2 | 0.58 | 0.44 0.71 |
| Crackles/Rales | 8 | 0 38 | 88 | 77 94 | 11 | 0 48 | 84 | 73 92 | 0.8 | 0.1 4.9 | 1.0 | 0.9 1.3 | 0.48 | 0.39 0.57 |

Inter rater agreement measured using Gwet’s AC1 between the bedside physician who performed the point of care lung ultrasound and the over-reading physician using only archives was 68%. Most characteristics showed acceptable inter-rater reliability between the bedside read and images over read. Excess short non-coalescent B-lines and pleural thickening showed poor agreement likely reflecting both the subjectivity of these items and the difference between reviewing saved and real-time images.

**Table 3.** Inter rater agreement between a blinded over read relying only on saved images and the bedside interpretation of the treating physician. CI, confidence interval. Where the readings differed the bedside physician ultrasonographer interpretation was used.

| Ultrasound finding | % Agreement | 95% CI | Gwet AC1 | 95% CI |
| --- | --- | --- | --- | --- |
| Normal study | 71 | 59 82 | 0.44 | 0.22 0.66 |
| Excess Coalescent (long) B lines | 75 | 65 85 | 0.51 | 0.31 0.71 |
| Excess short B lines (comet tail) | 55 | 43 66 | 0.15 | -0.10 0.39 |
| Effusion | 91 | 84 97 | 0.90 | 0.81 0.98 |
| Air bronchograms | 69 | 58 79 | 0.51 | 0.31 0.72 |
| Thickened/moth-eaten pleura | 53 | 42 65 | 0.11 | -0.13 0.35 |
| Atelectasis | 69 | 58 79 | 0.51 | 0.31 0.71 |
| Consolidation | 80 | 71 90 | 0.74 | 0.60 0.88 |

PCR testing for SARS-CoV-2 was not always available and when it was a variety of tests performed at different sites were used. The results are shown in Table 4.

**Table 4.**
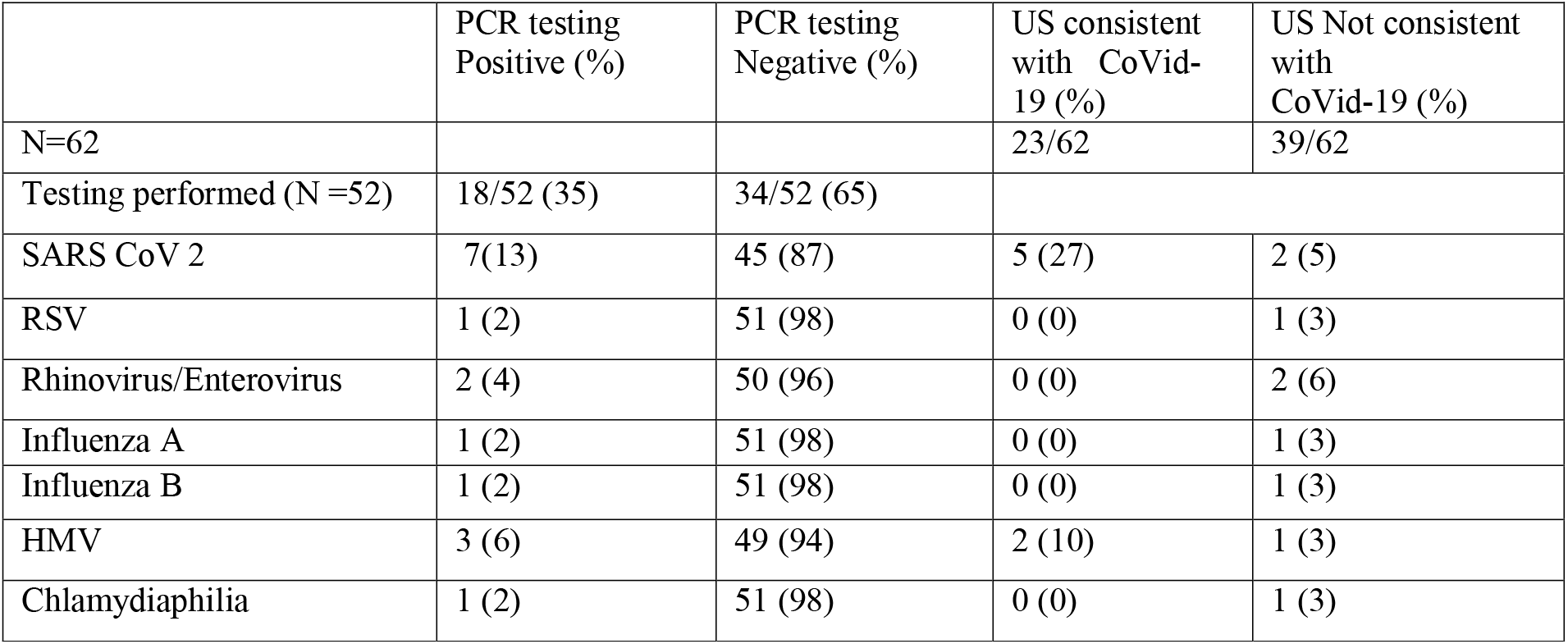
PCR results from nasal, nasopharyngeal, and or oropharyngeal swabs. Although the overall number of PCR tests was the same, some patients received SARS-CoV-2 testing alone, others had a panel of respiratory pathogens ordered without SARS-CoV-2 due to lack of test availability at the time. PCR, polymerase chain reaction; the panel of respiratory pathogens tested included Adenovirus, Parainfluenza viruses 1-4, Mycoplasma pneumoniae, Bordetella pertussis, Coronaviruses 229E, HKU1, N163 and OC43, RSV, respiratory syncytial virus; HMV, human metapneumovirus; Chlamydiaphilia, Chlamydiaphilia pneumoniae; US, ultrasound; HMV, human metapneumovirus.

## Discussion

Point of care lung ultrasound detected lesions consistent with alveolar involvement in 33% of symptomatic patients with normal vital signs who were being screened for CoVid-19.

These findings are consistent with published case series and social media reports of the utility of lung ultrasound in the diagnosis of CoVid-19.[19, 20] The use of point of care lung ultrasound in Covid-19 screening has been spontaneous and sporadic practice tending typically occurring in emergency medicine and critical care. Some radiologists have also found lung ultrasound useful.[19, 20] Regardless of the specialty, point of care lung ultrasound practices in the detection of CoVid-19 have necessarily evolved ahead of their published evidence base. The peer reviewed literature is sparse. Previous literature has comprised of case reports, and case series of 12 and 20 patients.[20, 21],[22] Scanning techniques, and images of patients with proven Covid-19 have spread among clinicians on Twitter and blogs[23, 24] and many others.

Our results may not be viewed so favorably to those who argue for implementation of CT scanning as the initial screening test. A CT only imaging approach by definition, will lead to better detection rates among those images than LUS. Such an approach, however, is expensive and slow and places a greater number of hospital staff at risk of infection, although it results in less infected patient exposure time for the doctor. It is possible that dedicated portable chest CT scanners using three slice protocols to save time and radiation would be a better alternative. This approach would require resources that are much less widely available than ED ultrasound.

Lung ultrasound has emerged as a clinical tool in human and veterinary medicine, and animal research with some advocates calling for it to replace the stethoscope. [25–28] Others have shown ultrasound to complement rather than replace the physical exam and to correlate reasonably well with lung findings at necropsy. Ultrasound decreases CT utilization in inpatients with suspected CoVid-19.[29] Descriptive papers have found ultrasound correlates well with CT and clinical characteristics in CoVid-19 patients. [30, 31] Recommendations for training novices to identify CoVid-19 have been published.[29] Ultrasound cannot be expected to replace CT imaging, but the ease with which it can be performed serially and at the bedside, and its apparent sensitivity makes it a useful tool for screening for alveolar level disease in SARS-CoV-2 infection.

Knowing whether a patient has alveolar involvement with CoVid-19 is clinically important. Patients’ initially mild lung disease has been shown to progress, sometimes rapidly, on serial CT scans as the disease progresses. [32] Patients frequently are unaware of their own deterioration and may present, or fail to re-present with critically low oxygen saturations without overt symptoms. These patients frequently have negative PCR tests unless bronchoalveolar lavage is performed. Such patients risk being falsely reassured about their own impending fate, and continue to infect others when, inevitably, they cough.

## Limitations

This was a single center study and was not a random sample. Whether a patient was seen by a physician who both believed that CoVid-19 screening should include lung imaging and was facile with ultrasound was a matter of luck, but not randomization. This adds uncertainty to estimates of the prevalence of pneumonitis that point of care lung ultrasound can detect among patients being screened for CoVid-19. Other limitations of our work include its small sample size, and a single chart reviewer. Patients with mild disease, and especially those with normal vital signs did not always have CT imaging performed. PCR testing for SARS-CoV-2 was not always available and even when it was the ideal gold standard of bronchoalveolar lavage was not performed.

Our use of CT scan as a gold standard is imperfect as CT diagnosis of CoVid-19 has its own limitations.[33] It is difficult to conceive of an alternative gold standard that does not fall foul of circular reasoning (by for example using ‘two out of three’ imaging methods positive as the gold standard). Moreover, given the false negative rates of PCR testing, CT rather than PCR testing has been recommended as the primary diagnostic modality in high prevalence settings. [34]

Our assessment of the performance characteristics of ultrasound is limited by our sample size. The performance characteristics of lung ultrasound were not the primary concern of this manuscript however. Although falling out of favor, null hypothesis testing is well suited to answering our primary question when the sample size is small– after all, single brown (red) Holstein demolishes the hypothesis that all cows are black and white, and careful planning minimizes the number of cows that need to be seen.

Despite these limitations, we can be assured that the prevalence of pneumonitis in these patients is more than the 2% ‘acceptable miss rate’ for high morbidity conditions and this may be sufficient to adjust practice accordingly. [35] Other limitations include the use of abbreviated LUS imaging protocols and the variability in image saving practices with doctors saving many cine-clips, and others only one or two still images. These differences in practice style could decrease inter rater agreement between the blinded and bedside readings. Much more detailed and formalized lung ultrasound protocols and ultrasound scoring systems specifically for use in SARS-CoV-2 patients have been described.[31, 36] Abbreviated protocols are inevitable in community practice and could lead to missed diagnoses. This would bias our study in the opposite direction of our actual findings.

## Conclusion

In conclusion, point of care lung ultrasound has a meaningful detection rate for alveolar level involvement in SARS-CoV-2 infection when screening symptomatic emergency department patients with normal vital signs. We recommend at least point of care lung ultrasound be used in addition to PCR testing when screening symptomatic patients in whom SARS-CoV-2 infection is suspected.

## Data Availability

Statistical code used in the analysis is available but the data is not but a public use dataset may be released with a peer-reviewed publication.

## Acknowledgements

The authors acknowledge Kimberly Kolbias, DO, a visiting resident from Desert Regional Medical Center, who posed the question “Do I really need to do the ultrasound if the vital signs are normal?” “Yes, you do.” This project was supported in part by the Pediatric Emergency Medicine Research Foundation, (Walsh) and by The National Institutes of Health through grant UL1 TR001860 (Bang). The ultrasound machine was provided by The Sutter Medical Foundation.

